# The inflammatory marker glycoprotein acetyls is associated with blood pressure and pulse wave velocity through childhood and adulthood: analysis of three population-based longitudinal cohorts

**DOI:** 10.64898/2025.12.01.25341421

**Authors:** Toby Mansell, Richard Liu, Joel Nuotio, Gabrielle PD MacKechnie, Katherine Lange, Mengjiao Liu, Fiona Mensah, Mika Ala-Korpela, Olli T Raitakari, Terho Lehtimäki, Mika Kähönen, Richard Saffery, Melissa Wake, Markus Juonala, Deborah A Lawlor, Neil Goulding, David Burgner

## Abstract

**Background:** Chronic inflammation is associated with the development of atherosclerosis across the life course, but the effect of inflammation on preclinical cardiovascular measures is poorly characterized, particularly in children and younger adults. We investigated cross-sectional associations of inflammatory markers with preclinical cardiovascular measures in three population-based cohorts of children and adults.

**Methods:** We analyzed data from 9865 children and adolescents and 11,086 adults from: the Longitudinal Study of Australian Children’s Child Health CheckPoint (CheckPoint, Australia) (1325 parents, mean age 44.6 years; 1180 offspring, mean age 12.0 years), the Avon Longitudinal Study of Parents and Children (ALSPAC, UK) (6395 parents, mean age 49.2 years; 7371 offspring assessed at up to 3 time points, approximately 8, 18 and 24 years), and the Cardiovascular Risk in Young Finns Study (YFS, Finland) (2015, mean age 37.7 years). Inflammatory markers were glycoprotein acetyls (GlycA) and high sensitivity C-reactive protein (hsCRP). Cardiovascular measures included systolic (SBP) and diastolic blood pressure (DBP), carotid-femoral pulse wave velocity (PWV), and carotid intima-media thickness (cIMT).

**Results:** In confounder-adjusted models, higher GlycA was cross-sectionally associated with higher blood pressure and PWV across CheckPoint and ALSPAC parents and offspring at each time point (e.g., for ALSPAC parents, difference in mean SBP 1.00 mmHg per 100 µmol/L GlycA (95%CI: 0.69, 1.31), DBP 0.63 mmHg (0.43, 0.84); for CheckPoint parents, difference in mean PWV 0.16 m/s (0.09, 0.24)), and to a limited extent in YFS adults (difference in mean SBP 0.29 mmHg (0.02, 0.56), DBP 0.18 mmHg (-0.05, 0.40), PWV 0.01 m/s (-0.04, 0.05)). Higher hsCRP was also associated with higher blood pressure and PWV in some cohorts (e.g, in ALSPAC parents, difference in mean SBP 0.64 mmHg per 1 standard deviation increase in log-10 hsCRP (95%CI: 0.35, 0.92), DBP 0.93 mmHg (0.41, 1.45); in CheckPoint parents, PWV 0.7 m/s (0.0, 1.4)). Neither GlycA nor hsCRP were associated with consistent differences in cIMT, with evidence for an effect of higher GlycA and hsCRP on lower cIMT in some cohorts.

**Conclusions:** Higher inflammation may contribute to higher blood pressure and arterial stiffness (PWV) from childhood onwards. GlycA is more evidently associated with these vascular measures in childhood than hsCRP.

**Clinical Perspective:** *What is new?:*

- The association of inflammation with the development of cardiovascular disease risk-associated phenotypes across the life course is poorly understood at younger ages, despite adult data highlighting strong associations between inflammation and cardiovascular disease pathogenesis.
- High-sensitivity C-reactive protein (hsCRP) is the most widely investigated inflammation marker, but glycoprotein acetyls (GlycA) may be a more informative chronic subclinical inflammation marker for cardiovascular disease risk, including earlier in life.
- This study investigated the effect of inflammation measured by hsCRP and GlycA on key preclinical cardiovascular measures across childhood and adulthood in large international cohorts.

*What are the clinical implications?:*

- In cross-sectional analyses in different populations, higher GlycA is associated with higher blood pressure and pulse wave velocity, but not carotid intima-media thickness, from childhood to adulthood, after adjusting for traditional cardiovascular disease risk factors, with less consistent effects observed for hsCRP.
- Reducing chronic inflammation in childhood may improve preclinical cardiovascular measures, with the potential to reduce cardiovascular disease across the life course.
- Additional analyses with longitudinal repeat measures and causal analyses, such as Mendelian randomization at different ages, as well as biomarkers and functional measures of inflammation are needed to inform whether these findings reflect causal effects and provide targets for prevention.

## Introduction

Atherosclerosis develops from early life onwards with a long preclinical period through childhood, adolescence and early to mid-adulthood^1^. This preclinical period is a largely over-looked opportunity for primordial and primary prevention before cardiovascular disease (CVD) is manifest and when pathological changes may be potentially reversible^1^.

Cumulative exposure to traditional cardiovascular risk factors (such as cigarette smoke exposure, obesity, diabetes, and dyslipidemia) in childhood and adolescence is associated with heightened inflammation^2^ and with higher risk of CVD events in adulthood^3^. Inflammation is implicated in all pathogenic stages of atherosclerosis, from foam cell infiltration of the arterial wall, through lesion development and maturation, to plaque rupture^4^. In adults with previous CVD events, some evidence supports targeting inflammation with either specific anti-cytokine biologics^5^ or non-specific anti-inflammatory therapies^6^ to reduce the incidence of further CVD events, independent of lipid levels. However, previous trials have predominantly involved men, and meta-analyses have used composite outcomes combining adverse cardiovascular event types due to low power for estimating effects on specific event types^6^.

Inflammatory biomarkers are also associated with preclinical cardiovascular measures such as blood pressure (BP)^7^, pulse wave velocity (PWV)^8^, and carotid intima-media thickness (cIMT)^9^. These preclinical measures are predictive of subsequent CVD events in adults^10^. Understanding the effects of inflammation on preclinical cardiovascular measures across childhood and early- and mid-adulthood would identify opportunities for primordial and primary prevention of CVD.

Glycoprotein acetyls (GlycA) is a nuclear magnetic resonance (NMR)-derived composite measure that is suggested to be more informative of cumulative inflammatory burden and a better predictor of CVD than the more widely used high-sensitivity C-reactive protein (hsCRP)^11^, with evidence for GlycA and hsCRP differentially associating with different types of CVD (myocardial infarction and ischemic stroke, respectively)^12^. The limited existing data in children and adults suggest there are effects of inflammation measured by GlycA and hsCRP on preclinical vascular measures^2,13–15^, but little is known about how these relationships change across the life course from childhood into early and mid-adulthood.

We therefore aimed to estimate the effect of chronic subclinical inflammation, measured by GlycA or hsCRP, on cross-sectional preclinical cardiovascular measures (BP, PWV, and cIMT) in children and adults in three large independent population-derived cohorts without prevalent CVD: the Longitudinal Study of Australian Children’s Child Health CheckPoint (CheckPoint), Avon Longitudinal Study of Parents and Children (ALSPAC) and Cardiovascular Risk in Young Finns Study (YFS).

## Methods

### Study designs and participants

We undertook cross-sectional analyses of adults and children from three cohorts that were selected as they had measures of GlycA, hsCRP and preclinical cardiovascular phenotypes in children and young adults. Within each cohort we selected which data collection phases to use based on maximising the sample size with available data and having a range of different ages in the adults and children (**Figure 1**).

**Figure 1.**
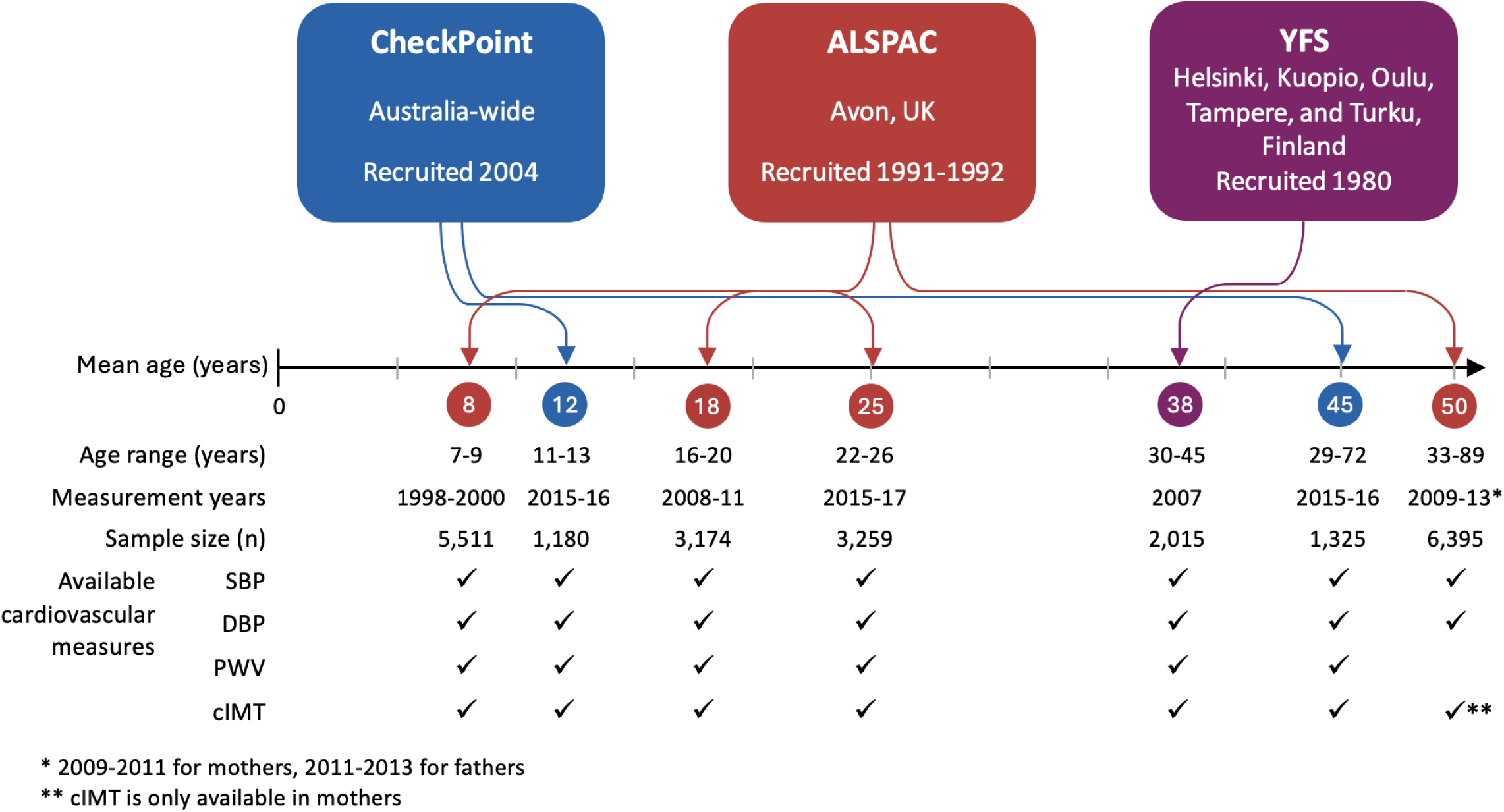
Summary of participants and data from each cohort used in this study.

#### Longitudinal Study of Australian Children’s Child Health CheckPoint

The Longitudinal Study of Australian Children (LSAC) recruited a nationally representative sample of 5107 infants (of 8921 families invited to participate, 57.2% participation rate) in 2004 using a 2-stage random sampling design with postcode as the primary sampling unit^16^. These children have subsequently been followed in ’waves’ of data collection every two years. Of the 3764 families retained in LSAC wave 6 in 2014, 3513 families consented to being contacted and were invited to undertake a detailed cross-sectional biophysical assessment (the Child Health CheckPoint, ‘CheckPoint’); 1874 families participated^16^ (53.3% of families invited to participate in CheckPoint, 36.7% of families originally recruited to LSAC). The child and a parent/caregiver (hereafter referred to as parents) attended one of 15 CheckPoint pop-up centres across Australia between February 2015 and March 2016. Inflammation and cardiovascular measures were available in 1180 children (33.6% of participants invited to CheckPoint, 23.1% of those initially recruited to LSAC), with mean age 12.0 years (range 11-13 years), and 1325 parents with mean age 44.6 years (range 29-72 years). Ethics approval for the study was obtained from The Royal Children’s Hospital (Melbourne, Australia) Human Research Ethics Committee (33225D) and The Australian Institute of Family Studies Ethics Committee (14-26).

#### Avon Longitudinal Study of Parents and Children (ALSPAC)

ALSPAC is a prospective population-based birth cohort study^17,18^, recruited from the former county of Avon in south-west England. Pregnant women with expected dates of delivery between April 1991 and December 1992 were invited to take part in the study. The initial number of pregnancies enrolled was 14,541 (of 20,248 eligible pregnancies, 71.8% particpation rate), resulting in 14,062 live births and 13,988 children who were alive at 1 year of age. Subsequent to this first phase of recruitment, additional eligible pregnancies and children were recruited across further phases of recruitment. The total sample size for analyses using any data collected after the age of seven is therefore 15,447 pregnancies, of which 14,901 children were alive at 1 year of age. 12,113 partners have been in contact with the study and 3,807 partners are currently enrolled.

For this study, we used data from 6395 parents (of 4969 offspring, 32.2% of recruited pregnancies), mean age 49.5 years, range 33-89 years) assessed in 2009-2011 for mothers and 2011-2013 for partners. We used data from three of the research clinic follow-up assessments of ALSPAC offspring. The first was undertaken 1998-2000, including 5511 participants (37.0% of original cohort [children alive at 1 year of age]) when they were mean age 7.5 years (range 7 to 9 years). The second was undertaken 2008-2011, including 3174 participants (21.3% of original cohort) when they were mean age 17.8 years (range 16 to 20 years). The third was undertaken 2015-2017, including 3259 (21.9% of original cohort) participants when they were mean age 24.5 years (range 22 to 26 years). Ethical approval for the study was obtained from the ALSPAC Ethics and Law Committee and the Local Research Ethics Committees.

### Cardiovascular Risk in Young Finns Study

The prospective population-based Cardiovascular Risk in Young Finns Study (YFS) was launched in 1980, with 3596 participants (of 4320 invited, 83.1% participation rate) aged 3 to 18 years randomly selected from the population register of the five Finnish university cities and their surrounding region. Participants have been assessed approximately every 3 years, as previously described^19^. For this study, we used data from 2015 participants (56.0% of the original cohort) in the 2007 follow-up study with available inflammation and cardiovascular measurements (mean age 37.7 years, range 30-45 years). Ethics approval for the study was obtained from the ethics committees of each participating university (Helsinki, Kuopio, Oulu, Tampere, and Turku).

Detailed information on each cohort, including participant consent, participant flow from recruitment to inclusion in each analysis (**Supplementary Figures S1-S5**), and comparisons of the distribution of maternal/sociodemographic characteristics between included participants and the original recruited cohorts (**Supplementary Table S1**), are presented in the supplementary material.

### Inflammation biomarkers

All blood samples were processed within 4 hours of collection with serum or plasma aliquots stored at -80°C. GlycA was measured in serum (CheckPoint^20^ and YFS) or EDTA-plasma (ALSPAC^21^) using a high-throughput proton NMR metabolomics platform (Nightingale Health, Vantaa, Finland). The NMR platform, quality control and applications have previously been described^22^. In ALSPAC mothers, GlycA was measured at two time points: between December 2008 and July 2011 (mean age 47 years [SD=4.5]), and between July 2011 and June 2013 (mean age 50 years [SD=4.4]). In the current analysis, we predominantly used GlycA measurements from the first assessment (47 years) but used data from the second (50 years) when a measurement was unavailable at the first assessment (n=217). The NMR platform also included several biomarkers relating to lipoproteins and other metabolites, and total triglycerides was included in the fully adjusted models due to the potential confounding of triglyceride levels in associations between inflammation and cardiovascular risk^23^. For all cohorts, the 2016 Nightingale Health bioinformatics pipeline was used^22^. High-sensitivity CRP levels were measured with the COBAS Integra Analyser (Roche Dignostics, NSW, Australia for CheckPoint; Roche UK, Welwyn Garden City, UK for ALSPAC) or auto-analyser Olympus AU400 (YFS). For ALSPAC children, hsCRP measurements from age 9-10 years were used as a proxy measure of hsCRP at 7-8 years.

### Cardiovascular measures

#### Blood pressure

Systolic and diastolic BP was measured using SphygmoCor XCEL (AtCor Medical Pty Ltd) for CheckPoint children and parents in supine, with the mean of at least two valid measurements used for analyses^24^. Omron BP/pulse monitor (Omron Healthcare Inc) was used to measure BP for ALSPAC parents and offspring at the 24-year-old time point and the Dinamap 9301 Vital Signs Monitor (Morton Medical) were used for offspring at the 7-year and 17-year time points. Measurements were taken while seated, and the mean of at least two measurements used for analyses. Random-zero sphygmomanometer (Hawksley & Sons Ltd) was used for YFS adults while seated, with the mean of three measurements used for analyses.

#### Carotid-femoral pulse wave velocity

In CheckPoint children and parents, carotid-femoral PWV was measured by SphygmoCor XCEL (AtCor Medical, Australia) as previously described and validated^24^. Following a seven-minute rest, assessors obtained velocity (distance/time) measurements one to three times with participants supine. Distance was measured from the carotid pulse to the suprasternal notch to the right femoral pulse (estimated by the crease between thigh and torso when the knee was bent to 90 degrees) to the top of the thigh cuff.

For the ALSPAC offspring, PWV was measured at 10-, 17- and 24-year time points. PWV was assessed at age 10 years using applanation tonometry, where pressure-pulse waveforms were recorded transcutaneously using a high-fidelity micro-manometer (SPC-301; Millar Instruments, Houston, TX) from the radial and carotid pulse synchronous with the ECG signal^25^, and used as a proxy measure in the 7-year time point analyses. For the 17-year and 24-year clinic assessment, a tonometer was placed over the right carotid artery in the participant’s neck, while another was located over the femoral artery in their upper right thigh. The distance between the participant’s suprasternal notch and the top of the thigh cuff was measured, as was the distance between their suprasternal notch and the bottom of the neck cuff on the right side. The equipment used in the PWV measurement consisted of a Vicorder instrument (Skidmore Medical) with two BP measurement channels and two Velcro pressure sensor cuffs (neck and leg). PWV was not measured in ALSPAC parents.

In YFS, a whole-body impedance cardiography device (CircMon; JR Medical, Ltd) was used to determine PWV. The CircMon software measures the time difference between the onset of the decrease in the whole-body impedance signal and the distal plethysmographic signal from a popliteal artery at the knee. A detailed description of the method and a validation study have been reported previously^26^, indicating that the whole-body impedance cardiography method had excellent reproducibility and good agreement with other methods such as Doppler ultrasound, though the impedance-based method overestimated PWV compared these other methods.

#### Carotid intima-media thickness

In CheckPoint children and parents, cIMT was measured as previously described^27^. Participants lay supine with their head turned 45 degrees to the left to expose the right side of neck. A 10 MHz linear array probe (Vivid-I, GE Healthcare, Chicago, IL, USA) captured cine-loops of the right common carotid artery. Maximum cIMT, calculated as the mean of 3-5 still frames timed at the R-wave by ECG, of the largest cIMT measurement in a 10 mm window proximal to the carotid bulb was used.

ALSPAC mothers (but not their partners) and their offspring at age 17 and 24 years (but not at 7 or 10 years) had cIMT measured with a 5-10 MHz linear array probe as described previously^28^. cIMT measurements were acquired from both the left and right common carotid artery arteries. The mean of the left- and right-sided readings was used in all analyses.

In YFS, ultrasound studies were performed using Sequoia 512 ultrasound mainframes (Acuson, CA, USA) with 13 MHz linear array transducers as previously described^29^. cIMT of the posterior (far) wall of the left carotid artery was measured.

### Other CVD risk factors and covariates

Covariates for inclusion in the full adjustment model as potential confounding factors were selected *a priori* based on content knowledge and availability of data for harmonised adjustment of models across all cohorts. All cohorts collected data on self- or parent-reported sex assigned at birth, and BMI was calculated as kg/m^2^. Height was measured to the nearest 0.1 cm (CheckPoint and ALSPAC) or 1 cm (YFS) and weight to the nearest 0.1 kg. For children, US Centers for Disease Control and Prevention (CDC) growth reference charts provided age- and sex-specific BMI z-scores and percentiles^30^.

Information on smoking and socioeconomic position (SEP) were obtained from questionnaires. In CheckPoint, for adults, current smoking status was determined by asking “are you currently smoking?” in the LSAC wave 6 questionnaire. Adults who indicated they had smoked in the past or answered yes to the smoking question at least once in previous LSAC waves, were considered ever-smokers. In children, a binary passive smoking exposure was derived from the parent response to the question “Including yourself, how many people who live with you smoke inside the house?” in each LSAC wave. Children were defined as exposed if the answer to this question was more than 0 at any wave, otherwise they were considered unexposed. The family SEP indicator used was highest level of parent education reported at LSAC wave 6, categorized into three groups: university degree or higher, other post-secondary school qualification, or secondary school completion or lower. Ethnicity was dichotomised based on parental country of birth and Indigenous or Torres Strait Islander status due to low numbers of participants of ethnic minority, with the participant being considered as being in an ethnic minority or mixed group if either parent was an Indigenous or Torres Strait Islander or born in a non-English speaking country, otherwise they were considered not to be in an ethnic minority group^31^. Puberty stage was measured by self-reported questionnaires with pictures of Tanner pubertal stages^32^.

For ALSPAC, current smoking status was assessed from questionnaires for mothers, young adults and teenagers, at a similar age to when GlycA, PWV and cIMT were measured; where this was not available, we used current smoking status from the most recent previous questionnaire. For participants without any data on current smoking, we categorized them as ever- or never-smoker.

Family SEP was assessed as the highest level of maternal/paternal education at the time of enrolment, categorized into 3 groups: university degree or above, A-level (Advanced Level; exams taken in different subjects usually at age 18 years), or below A-level. Ethnicity of mother and offspring was reported by mothers in questionnaires, dichotomized here as white or non-white due to low numbers of non-white participants.

In YFS, current smoking was defined as positive if participant smoked daily. Participants who indicated they had smoked daily for at least a year in the past were considered ever-smokers. Participants’ SEP was defined as the number of completed school years reported at the time of the 2007 follow-up study. Where these data were unavailable from the 2007 study (33.8% of participants), data from 2011 (31.6%) or 2001 (2.2%) was used instead. The YFS sample is considered racially homogenous, so ethnicity was not included in YFS analyses.

### Statistical analysis

We pre-specified confounders for the associations between exposures (inflammatory biomarkers GlycA and hsCRP) and outcomes (BP, PWV and cIMT) of interest. Included confounders were age, sex, ethnicity (for CheckPoint and ALSPAC), SEP, smoking (for adults) or passive smoke exposure (for children), BMI, and total triglycerides. To assess how representative the samples used in this study (i.e., those with the required data) were of the originally recruited cohorts, we compared the distribution of maternal demographics (maternal age, ethnicity, SEP, and smoking status) between those included in this study with the original cohort at recruitment. As hsCRP had a markedly skewed distribution, it was log-10-transformed prior to further analysis. Pearson’s correlations between GlycA and hsCRP at each age were estimated. Multiple linear regression models were used to examine cross-sectional associations between GlycA or hsCRP and cardiovascular measures (BP, PWV, and cIMT) separately at each age in each cohort. All models were complete case analyses. The main model included all observed confounders and was used to estimate the effect of inflammation measured by GlycA or hsCRP. Results were reported for GlycA in units of 100 µmol/L and for hsCRP in standard deviation units on the log-10 scale. To facilitate shared axes in the figures, estimates for PWV were rescaled to cm/s units and estimates for cIMT were rescaled to 10 µm units for the figures. Estimates are reported in natural units in the text. In supplementary material, figures are presented with units of GlycA and cardiovascular measures rescaled to standard deviation units (**Supplementary Figures S6** and **S7**). We explored sex differences by repeating confounder-adjusted models in females only and males only, and tested whether accounting for sex-differences substantially improved the model with likelihood ratio tests comparing the confounder-adjusted models with and without a sex*inflammation exposure interaction term. As the pubertal status of Checkpoint children spanned from pre-pubertal to post-pubertal, and pubertal status is associated with inflammation levels^33^, we performed sensitivity analysis to investigate if additionally adjusting the CheckPoint children models for pubertal stage substantially changed estimated associations between inflammation and cardiovascular measures. For all ages and cohorts, minimally adjusted models adjusting for age and sex only are included as secondary analysis. These additional analyses are presented in the supplementary material.

## Results

### Cohort summaries

In total, inflammatory marker and cardiovascular data were available for 9865 children and adolescents (<18 years; 50.1% males) and 12,994 adults (>18 years; 32.4% males). By cohort, GlycA data were available for 1325 adults (mean age 44.6 years) and 1180 children (12.0 years) in CheckPoint; 6395 parents (49.5 years), 3259 young adults (24.5 years), 3174 teenagers (17.8 years) and 5511 children (7.5 years) in ALSPAC, and 2015 adults (37.5 years) in YFS. Of those with GlycA data, hsCRP data were available for 1313 adults and 1156 children in CheckPoint; 6176 parents, 2943 young adults, 3174 teenagers and 3441 children in ALSPAC, and 2015 adults in YFS. The characteristics of the study group, including mean GlycA and hsCRP levels and vascular outcomes, are outlined in **Table 1** and **Table 2**. The comparison of maternal demographic variables at the time of recruitment between those included in this study and full recruited cohort is shown in **Supplementary Table 1**, with the CheckPoint sample in this study having lower diversity in ethnicity and higher levels of maternal education compared to the original cohort, the YFS sample having higher rates of maternal smoking, and the ALSPAC sample having higher levels of education and lower smoking rates.

**Table 1.**
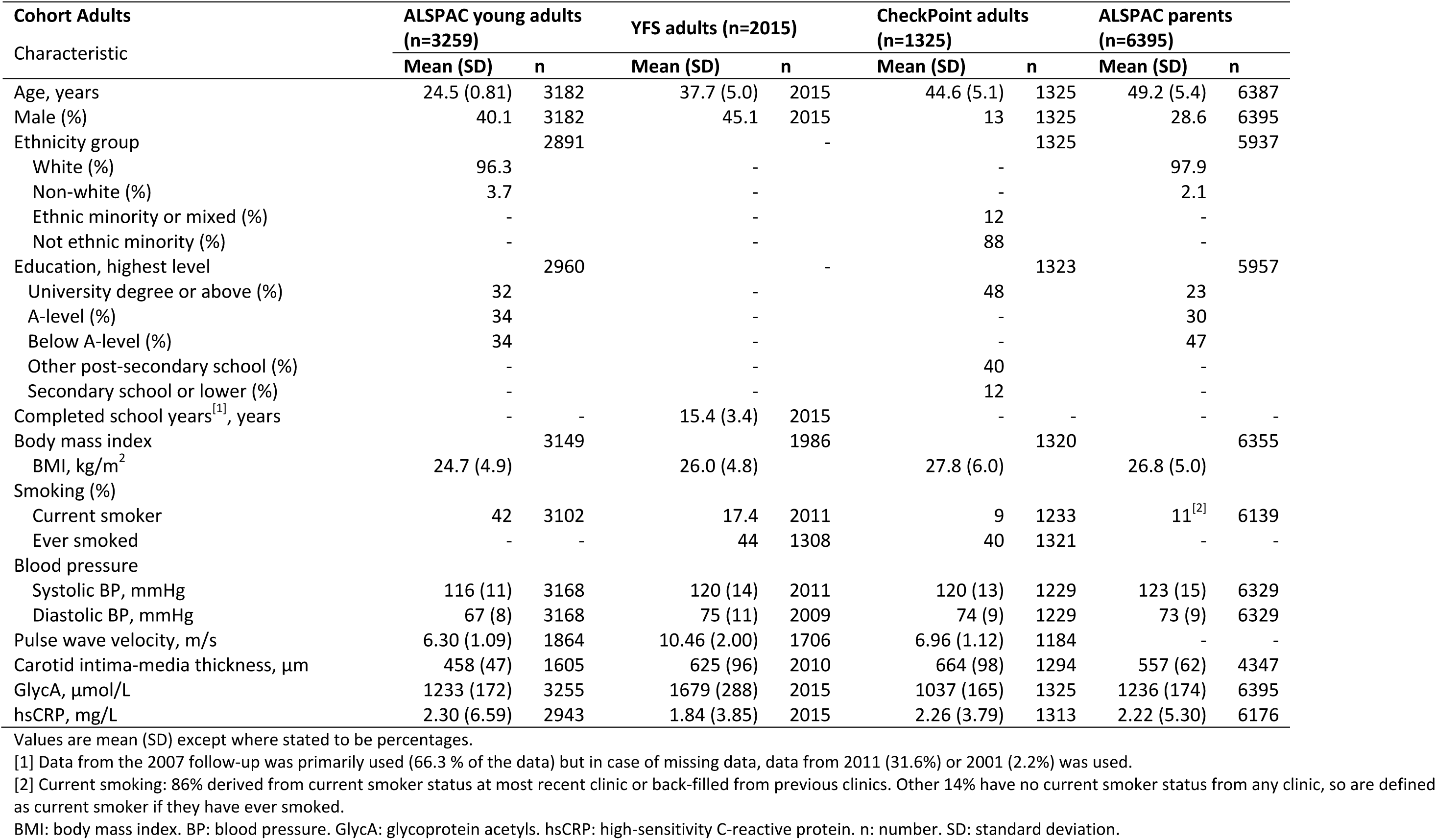
Baseline sample characteristics and data availability of adults who fulfil inclusion criteria (providing an analysable blood sample).

| Cohort Adults<br>Characteristic | ALSPAC young adults<br>(n=3259) |  | YFS adults (n=2015) |  | CheckPoint adults<br>(n=1325) |  | ALSPAC parents<br>(n=6395) |  |
| --- | --- | --- | --- | --- | --- | --- | --- | --- |
|  | Mean (SD) | n | Mean (SD) | n | Mean (SD) | n | Mean (SD) | n |
| Age, years | 24.5 (0.81) | 3182 | 37.7 (5.0) | 2015 | 44.6 (5.1) | 1325 | 49.2 (5.4) | 6387 |
| Male (%) | 40.1 | 3182 | 45.1 | 2015 | 13 | 1325 | 28.6 | 6395 |
| Ethnicity group |  | 2891 |  | - |  | 1325 |  | 5937 |
| White (%) | 96.3 |  | - |  | - |  | 97.9 |  |
| Non-white (%) | 3.7 |  | - |  | - |  | 2.1 |  |
| Ethnic minority or mixed (%) | - |  | - |  | 12 |  | - |  |
| Not ethnic minority (%) | - |  | - |  | 88 |  | - |  |
| Education, highest level |  | 2960 |  | - |  | 1323 |  | 5957 |
| University degree or above (%) | 32 |  | - |  | 48 |  | 23 |  |
| A-level (%) | 34 |  | - |  | - |  | 30 |  |
| Below A-level (%) | 34 |  | - |  | - |  | 47 |  |
| Other post-secondary school (%) | - |  | - |  | 40 |  | - |  |
| Secondary school or lower (%) | - |  | - |  | 12 |  | - |  |
| Completed school years<sup>[1]</sup>, years | - | - | 15.4 (3.4) | 2015 | - | - | - | - |
| Body mass index |  | 3149 |  | 1986 |  | 1320 |  | 6355 |
| BMI, kg/m<sup>2</sup> | 24.7 (4.9) |  | 26.0 (4.8) |  | 27.8 (6.0) |  | 26.8 (5.0) |  |
| Smoking (%) |  |  |  |  |  |  |  |  |
| Current smoker | 42 | 3102 | 17.4 | 2011 | 9 | 1233 | 11<sup>[2]</sup> | 6139 |
| Ever smoked | - | - | 44 | 1308 | 40 | 1321 | - | - |
| Blood pressure |  |  |  |  |  |  |  |  |
| Systolic BP, mmHg | 116 (11) | 3168 | 120 (14) | 2011 | 120 (13) | 1229 | 123 (15) | 6329 |
| Diastolic BP, mmHg | 67 (8) | 3168 | 75 (11) | 2009 | 74 (9) | 1229 | 73 (9) | 6329 |
| Pulse wave velocity, m/s | 6.30 (1.09) | 1864 | 10.46 (2.00) | 1706 | 6.96 (1.12) | 1184 | - | - |
| Carotid intima-media thickness, µm | 458 (47) | 1605 | 625 (96) | 2010 | 664 (98) | 1294 | 557 (62) | 4347 |
| GlycA, µmol/L | 1233 (172) | 3255 | 1679 (288) | 2015 | 1037 (165) | 1325 | 1236 (174) | 6395 |
| hsCRP, mg/L | 2.30 (6.59) | 2943 | 1.84 (3.85) | 2015 | 2.26 (3.79) | 1313 | 2.22 (5.30) | 6176 |

Values are mean (SD) except where stated to be percentages.
[1] Data from the 2007 follow-up was primarily used (66.3 % of the data) but in case of missing data, data from 2011 (31.6%) or 2001 (2.2%) was used.
[2] Current smoking: 86% derived from current smoker status at most recent clinic or back-filled from previous clinics. Other 14% have no current smoker status from any clinic, so are defined as current smoker if they have ever smoked.
BMI: body mass index. BP: blood pressure. GlycA: glycoprotein acetyls. hsCRP: high-sensitivity C-reactive protein. n: number. SD: standard deviation.

**Table 2.**
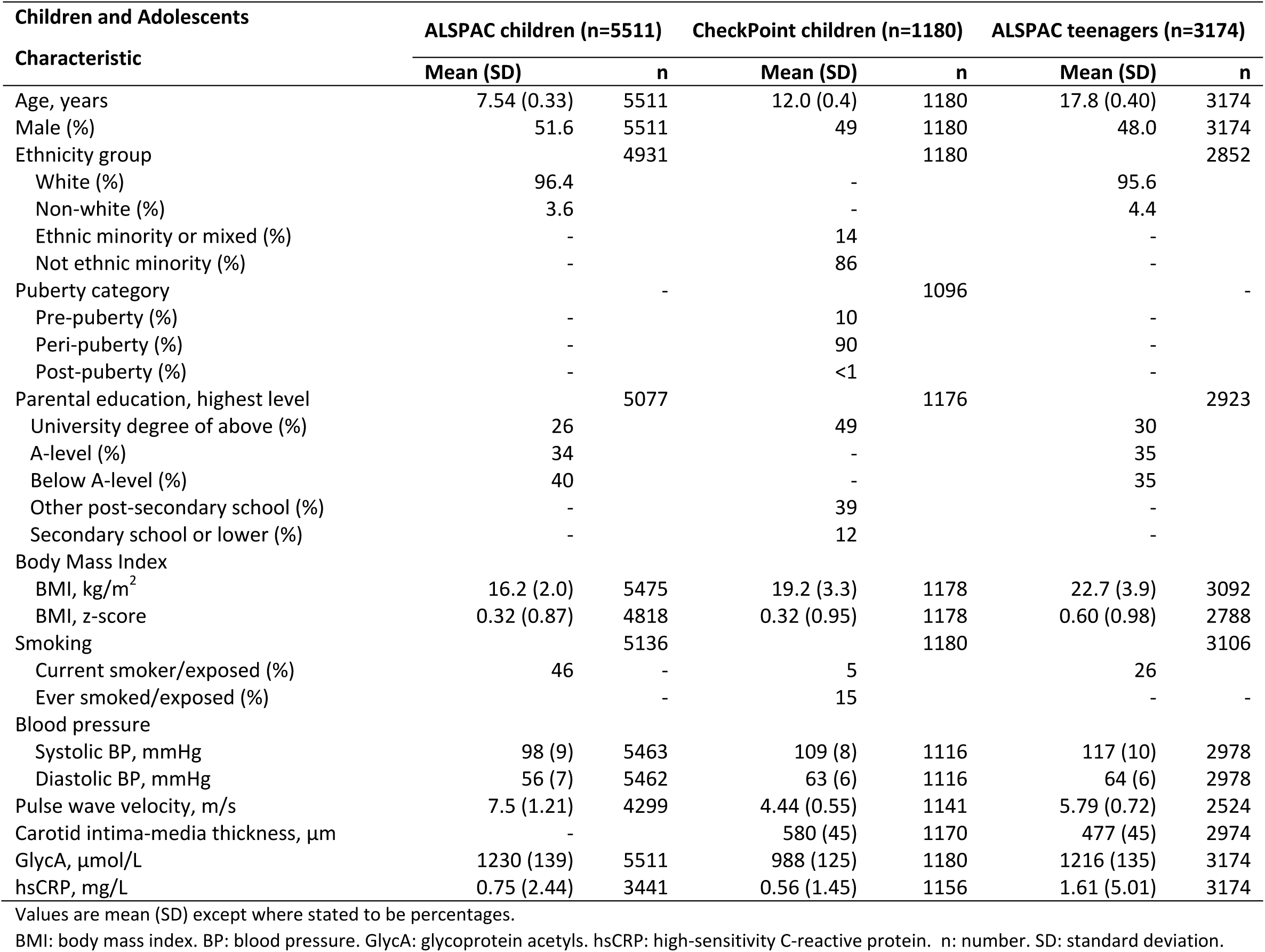
Baseline sample characteristics and data availability of children and teenagers who fulfil inclusion criteria (providing an analysable blood sample).

### Inflammation biomarkers and preclinical cardiovascular measures

The correlation between GlycA and hsCRP was lowest in CheckPoint and highest in ALSPAC (CheckPoint adults r=0.38, CheckPoint children r=0.23, YFS r=0.43, ALPSAC parents r=0.53, ALSPAC young adults r=0.55, ALSPAC teenagers r=0.59), with the exception of the ALSPAC children (r=0.17), where GlycA was measured at mean age 7 and hsCRP was measured at age 9. The associations between inflammation measured by GlycA or by hsCRP and each cardiovascular measure in the main models are shown in **Figure 2** and **Figure 3**, respectively. Higher GlycA was generally associated with higher systolic and diastolic BP across each age (e.g., difference in mean systolic BP ranged from 0.29 to 1.01 mmHg per 100 µmol/L GlycA), with the strongest association seen for CheckPoint adults and the weakest for YFS adults. For hsCRP, a similar pattern between higher hsCRP and higher systolic and diastolic BP was observed, particularly for the ALSPAC parents, young adults and teenagers, but the confidence interval for the estimated effect of hsCRP included the null for several of the ages.

**Figure 2.**
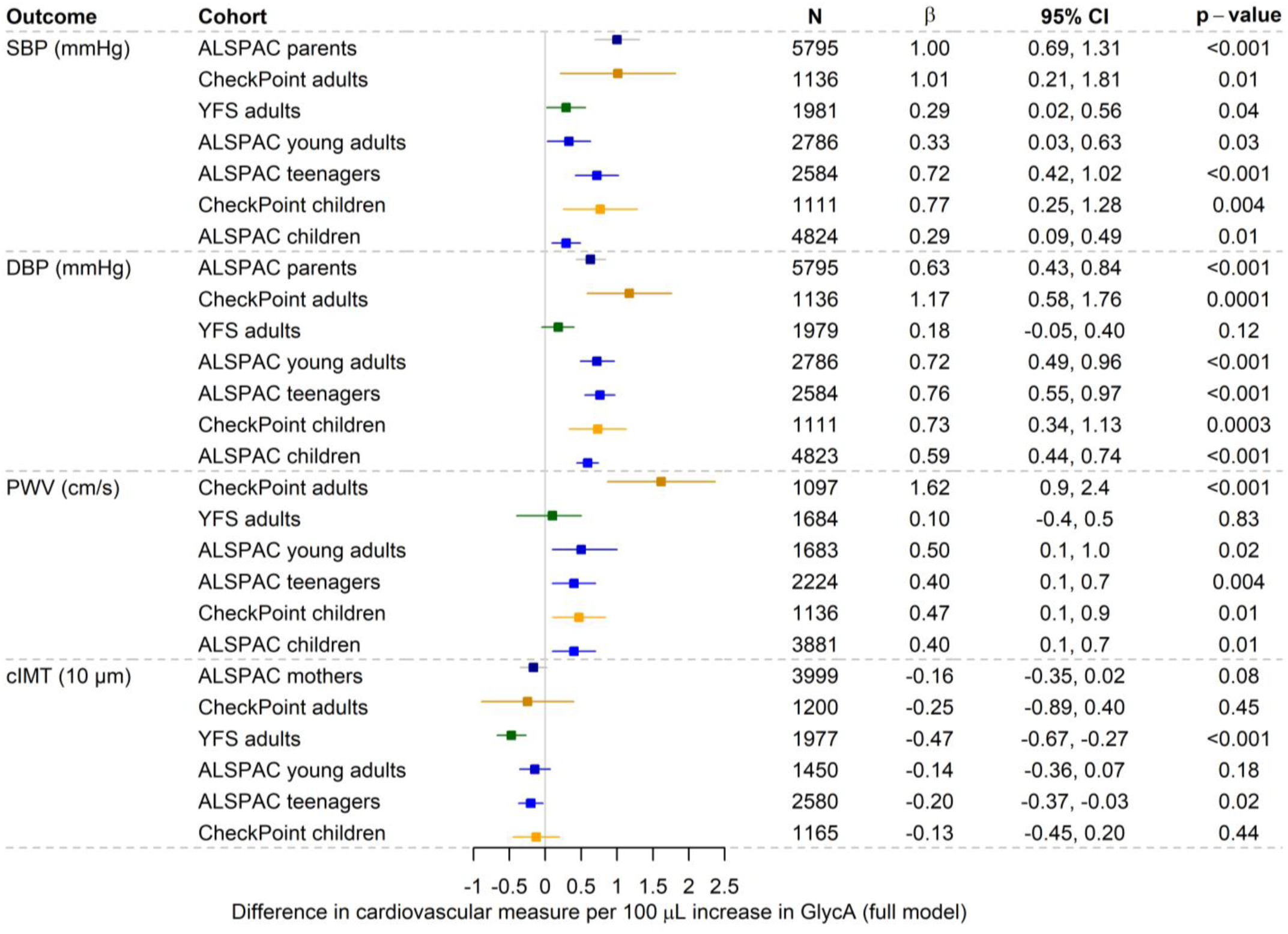
Association between GlycA and each preclinical cardiovascular measure for each time point/cohort. Estimates are reported in units specified for each outcome with GlycA units of 100 µL. The models were adjusted for age, sex, race/ethnicity (for CheckPoint and ALSPAC), SEP, smoking (for adults) or passive smoke exposure (for children), BMI, and total triglycerides. Bars are 95% confidence intervals. CI: confidence interval. cIMT: carotid intima-media thickness. DBP: diastolic blood pressure. GlycA: glycoprotein acetyls. PWV: pulse wave velocity. SBP: systolic blood pressure.

**Figure 3.**
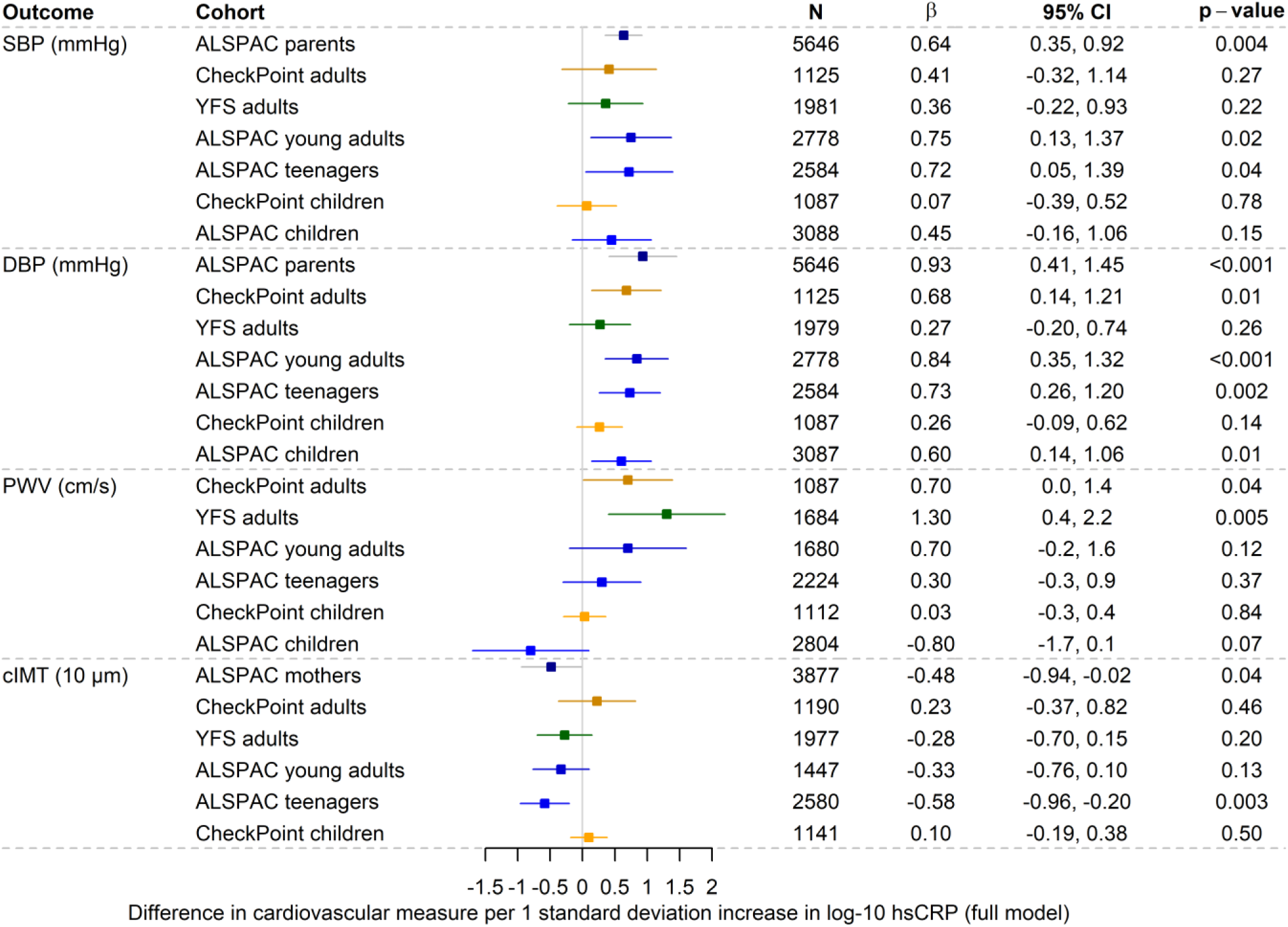
Association between hsCRP and each preclinical cardiovascular measure for each time point/cohort. Estimates are reported in units specified for each outcome per 1 standard deviation increase in log-10 hsCRP. The models were adjusted for age, sex, race/ethnicity (for CheckPoint and ALSPAC), SEP, smoking (for adults) or passive smoke exposure (for children), BMI, and total triglycerides. Bars are 95% confidence intervals. CI: confidence interval. cIMT: carotid intima-media thickness. DBP: diastolic blood pressure. hsCRP: high-sensitivity C-Reactive Protein. PWV: pulse wave velocity. SBP: systolic blood pressure.

Higher GlycA was associated with higher PWV for all ages (except for the YFS adults), with a larger association seen for CheckPoint adults (mean difference in PWV of 0.16 m/s per 100 µmol/L GlycA, 95% confidence interval [95% CI] 0.09 to 0.24, p<0.001) than the other ages. Higher hsCRP was mainly associated with higher PWV in the CheckPoint adults and YFS adults only (e.g., for YFS: 0.13 m/s per standard deviation unit of log-10 hsCRP, 95% CI 0.04 to 0.22, p=0.005).

In contrast to the expected direction of association, higher GlycA was associated with lower cIMT across some ages, particularly the YFS adults (mean difference in cIMT of −0.47 µm per 100 µmol/L GlycA, 95% CI −0.67 to −0.27). Higher hsCRP was also associated with lower cIMT across multiple ages, which was most evident in ALSPAC mothers and ALSPAC teenagers.

## Additional analyses

The association of GlycA and hsCRP with the preclinical cardiovascular measures in sex-stratified models are shown in **Supplementary Figure S8** and **Supplementary Figure S9**, respectively. There was statistical evidence for sex differences for some associations between inflammation and cardiovascular measures (CheckPoint adults for hsCRP with cIMT; ALSPAC young adults for GlycA with PWV, cIMT and SBP, and hsCRP with cIMT; YFS for hsCRP with PWV and cIMT) (**Supplementary Table S2**). For these models, inflammation was generally associated with larger differences in preclinical cardiovascular measures for females than males. Results with additional adjustment for puberty stage in CheckPoint children were consistent with those in the main analyses (**Supplementary Figure S10**). The associations between the inflammatory markers and preclinical cardiovascular measures were generally stronger in the minimally adjusted models compared to the main analyses (**Supplementary Figures S11** and **S12**).

## Discussion

We found higher inflammation measured by GlycA was consistently cross-sectionally associated with higher systolic and diastolic BP across population-based child and adult cohorts, and associated with higher PWV across most cohorts, after adjustment for traditional risk factors. hsCRP was also associated with higher BP and PWV, albeit less consistently, with less evidence of associations with PWV at younger ages. These findings suggest that GlycA may be a superior marker of inflammation-associated arterial stiffness than hsCRP from mid-childhood through mid-adulthood. Neither GlycA nor hsCRP were consistently associated with cIMT, with higher GlycA and hsCRP both associated with lower cIMT at some ages.

Levels of GlycA and hsCRP are moderately correlated with each other^34^, as also observed in this study, and these biomarkers are thought to capture overlapping but distinct inflammatory pathways^35^. GlycA and hsCRP are independently associated with CVD risk, even after mutual adjustment^34,35^. GlycA is a composite measure of glycosylation of several acute phase proteins and potentially reflects many inflammatory pathways. The process of glycosylation protects proteins from degradation or proteolysis and GlycA levels have higher stability over time compared to hsCRP^36^. GlycA has therefore been suggested to better reflect cumulative inflammation than hsCRP, which is predominantly an acute phase reactant, especially in children^37^. C-reactive protein is a single gene product, largely induced by IL-6, and is highly sensitive to acute inflammatory stimuli, such as infection, with higher intra-individual variability^37^.

We found that higher GlycA was generally associated with higher BP and PWV, while less consistent evidence was seen for cIMT, with a negative relationship between GlycA and cIMT seen for some ages. Higher blood pressure is a well-established risk factor for future CVD in adults, with considerable evidence for high blood pressure tracking from childhood into adulthood^38^. Both PWV and cIMT are also associated with risk of future CVD^10^. PWV, a measure of arterial stiffness, appears to enhance CVD prediction beyond traditional risk factors^39^, both in high-risk groups and healthy populations^40^. Carotid IMT has previously been described as a measure of subclinical atherosclerosis, although this is debated^41^ as it may be more reflective of arterial injury. Carotid IMT is not currently recommended for systematic use in CVD risk prediction by the European Society of Cardiology^42^, although this remains somewhat controversial^43^.

Previous studies, largely in adults, have investigated the association of GlycA and/or hsCRP with BP, PWV, or cIMT^44,45^. There are fewer analogous data in children^46,47^ and adolescents^48^, often from small and/or high-risk cohorts, such as individuals with severe obesity^48^. This is the first cross-cohort study that combines available data from three cohorts in harmonised models across different stages of life from childhood to adulthood. In adult studies that have considered both inflammatory markers, GlycA has generally showed a stronger association with preclinical cardiovascular measures than hsCRP in models adjusting for BMI^2,45^. This is consistent with our findings that GlycA had a more consistent relationship with BP and PWV than hsCRP, particularly at younger ages. Notably, while consistently positive associations were seen between GlycA and PWV across the CheckPoint and ALSPAC cohorts, this was not the case for YFS adults. This could be related to the differences in PWV measurement method, with CheckPoint and ALSPAC using applanation tonometry and YFS using whole-body impedance cardiography, which tends to overestimate PWV compared to other methods^26^, consistent with the higher PWV values we observed in YFS adults compared to CheckPoint and ALSPAC. Differences between cohorts may also reflect population-specific differences in determinants of cardiovascular risk more generally, as the association between GlycA and diastolic BP was lower in magnitude for YFS adults compared to the other cohorts, and there was also stronger evidence for the association between higher GlycA and lower cIMT in this cohort. For some associations, primarily between hsCRP and cIMT, there was evidence for sex differences with this association larger in females than males for several ages. Previously an observational study reported a larger association of GlycA and hsCRP with risk of ischemic stroke in males than females^12^, though participants in trials of anti-inflammatory interventions have predominantly been men^6^, and sex differences in treatment effects have not been robustly assessed to date.

A key strength of the current study is the harmonised analysis of three independent population-based cohorts in different settings with ages from childhood to mid-adulthood, allowing investigation of how patterns between these inflammation markers and preclinical cardiovascular measures may differ at different ages and in different settings. Limitations include the cross-sectional analysis of data in each cohort. While we hypothesised a causal effect of inflammation on these preclinical cardiovascular measures, the associations we have observed could be explained by factors such as residual confounding, reverse causality (i.e., preclinical cardiovascular traits influencing inflammatory markers), and selection bias resulting from loss to follow-up of participants and the complete case approach used for analyses. Studies considering longitudinal associations, and approaches such as Mendelian randomization^49^, are needed to build evidence for causal effect of inflammation on these preclinical cardiovascular measures, particularly earlier in the life course. Inflammation may offer a target for primordial and primary prevention of cardiovascular disease risk. Methodological differences between cohorts, including for the cardiovascular measures, and the differences in epoch of recruitment may contribute in part to the inter-cohort discrepancies in findings. There was modest reproducibility of cIMT measurements in CheckPoint ^27^, which may have biased estimated associations towards the null. In addition, CheckPoint and ALSPAC participants lacked ethnic and socioeconomic diversity (as used in this study) compared to the general populations from which they were drawn, a frequent limitation of cohorts from high-income settings^50^. The YFS participants are homogenous in terms of ethnicity, limiting generalizability to other populations. Findings should be replicated in other settings and populations, including those with a disproportionately high CVD burden, who may benefit most from primordial and primary prevention^1^.

## Conclusions

In this study of cross-sectional population-based analyses of mid-childhood to adulthood cohorts from three countries, inflammation was associated with adverse cardiovascular measures. Higher GlycA was consistently associated with BP at each age and with PWV at most ages. High sensitivity CRP was generally associated with BP, albeit less consistently and was only associated with higher PWV in older adults. These findings suggest chronic inflammation measured by GlycA is potentially more informative of CVD risk from childhood than inflammation measured by hsCRP, in keeping with analogous data from adults. Further studies are needed to understand the age-specific and longitudinal relationships between inflammation and CVD risk from childhood onwards and to identify upstream modifiable drivers of inflammation and functional targets for primordial and primary prevention earlier in life.

## Data Availability

Data used in this manuscript are available upon request through cohort-specific data access applications to the applicable cohort data custodians.

## Acknowledgements

### CheckPoint

This study uses data from the Longitudinal Study of Australian Children (LSAC) and Child Health CheckPoint. LSAC was conducted in partnership between the Department of Social Services, the Australian Institute of Family Studies and the Australian Bureau of Statistics (ABS), while the CheckPoint was led from the Murdoch Children’s Research Institute (MCRI). The findings and views reported in this paper are those of the author and should not be attributed to DSS, AIFS or the ABS. We thank the LSAC and CheckPoint study participants and families. We also thank the CheckPoint team and the MRCI.

### ALSPAC

We are extremely grateful to all the families who took part in this study, the midwives for their help in recruiting them, and the whole ALSPAC team, which includes data collection staff, data and administrations staff, technical managers and the technical staff with the Bristol Bioresource Laboratory, based within the University of Bristol.

### YFS

We thank the teams that collected data at all measurement time points; the persons who participated as both children and adults in these longitudinal studies; and biostatisticians Irina Lisinen, Johanna Ikonen, Noora Kartiosuo, Ville Aalto, and Jarno Kankaanranta for data management and statistical advice.

## Sources of Funding

### CheckPoint

This work has been supported to date by the National Health and Medical Research Council of Australia (NHMRC; 1041352, 1109355), The Royal Children’s Hospital Foundation (2014–241), Murdoch Children’s Research Institute, The University of Melbourne, National Heart Foundation of Australia (100660), Financial Markets Foundation for Children (2014–055; 2016–310) and Victoria Deaf Education Institute. Research at the Murdoch Children’s Research Institute is supported by the Victorian Government’s Operational Infrastructure Program. This paper uses unit record data from the Longitudinal Study of Australian Children. The study was conducted in partnership between the Department of Social Services (DSS), the Australian Institute of Family Studies (AIFS) and the Australian Bureau of Statistics (ABS). The findings and views reported in this paper are those of the authors and should not be attributed to DSS, AIFS or the ABS.

### ALSPAC

The UK Medical Research Council and Wellcome (Grant ref: MR/Z505924/1) and the University of Bristol provide core support for ALSPAC. This publication is the work of the authors and NG will serve as guarantor for the ALSPAC content of this paper. A comprehensive list of grants funding is available on the ALSPAC website (http://www.bristol.ac.uk/alspac/external/documents/grant-acknowledgements.pdf); This research was specifically funded by NIHR (Grant ref: NF-SI-0611-10196), MRC (Grant ref: MC_UU_12013/1), British Heart Foundation (Grant ref: CS/15/6/31468, SP/07/008/24066 and RG/10/004/28240). NG was supported by The John Templeton Foundation (Grant ref: 61917). The opinions expressed in this publication are those of the authors and do not necessarily reflect the views of the John Templeton Foundation.

### YFS

The Young Finns Study has been financially supported by the Academy of Finland: grants 356405, 322098, 286284, 134309 (Eye), 126925, 121584, 124282, 129378 (Salve), 117797 (Gendi), and 141071 (Skidi); the Social Insurance Institution of Finland; Competitive State Research Financing of the Expert Responsibility area of Kuopio, Tampere and Turku University Hospitals (grant X51001); Juho Vainio Foundation; Paavo Nurmi Foundation; Finnish Foundation for Cardiovascular Research; Finnish Cultural Foundation; The Sigrid Juselius Foundation; Tampere Tuberculosis Foundation; Emil Aaltonen Foundation; Yrjö Jahnsson Foundation; Signe and Ane Gyllenberg Foundation; Diabetes Research Foundation of Finnish Diabetes Association; EU Horizon 2020 (grant 755320 for TAXINOMISIS and grant 848146 for To Aition); European Research Council (grant 742927 for MULTIEPIGEN project); Tampere University Hospital Supporting Foundation; Finnish Society of Clinical Chemistry; the Cancer Foundation Finland; pBETTER4U_EU (Preventing obesity through Biologically and bEhaviorally Tailored inTERventions for you; project number: 101080117); CVDLink (EU grant nro. 101137278) and the Jane and Aatos Erkko Foundation. Pashupati P. Mishra was supported by the Academy of Finland (Grant number: 349708) and Emma Raitoharju (grants: 330809, 338395).

### Authors

T.M. is supported by a philanthropic fellowship from The DHB Foundation as managed by Equity Trustees. M.A.-K. was supported by a research grant from the Sigrid Juselius Foundation, the Finnish Foundation for Cardiovascular Research, and the Research Council of Finland (grant no. 357183). D.B. is supported by an NHMRC Investigator Grant (GTN1175744). DAL’s contributions are supported by the UK Medical Research Council (MC_UU_00032/05) and British Heart Foundation (CH/F/20/90003 & AA/18/1/34219). MW is supported by NHMRC Investigator Grant 2035040.

## Disclosures

The authors have no conflicts of interest to declare.

## Non-standard Abbreviations and Acronyms

ALSPAC-: Avon Longitudinal Study of Parents and Children
BP-: blood pressure
CDC-: US Centers for Disease Control and Prevention
CheckPoint-: Child Health CheckPoint
cIMT-: carotid intima-media thickness
CRP-C-: Reactive Protein
CVD-: cardiovascular disease
GlycA-: glycoprotein acetyls
hsCRP-: high-sensitivity C-Reactive Protein
IL-: Interleukin
LSAC-: Longitudinal Study of Australian Children
NMR-: nuclear magnetic resonance
PWV-: pulse wave velocity
SD-: standard deviation
SEP-: socioeconomic position
YFS-: Cardiovascular Risk in Young Finns Study

## Notes

### Competing Interest Statement

The authors have declared no competing interest.

### Author Declarations

CheckPoint: Ethics approval for the study was obtained from The Royal Children's Hospital (Melbourne, Australia) Human Research Ethics Committee (33225D) and The Australian Institute of Family Studies Ethics Committee (14-26). ALSPAC: Ethical approval for the study was obtained from the ALSPAC Ethics and Law Committee and the Local Research Ethics Committees. YFS: Ethics approval for the study was obtained from the ethics committees of each participating university (Helsinki, Kuopio, Oulu, Tampere, and Turku).

